# Genomic surveillance reveals early detection and transition of Delta to Omicron Lineages of SARS-CoV-2 Variants in wastewater treatment plants of Pune, India

**DOI:** 10.1101/2023.02.21.23286222

**Authors:** Vinay Rajput, Rinka Pramanik, Vinita Malik, Rakeshkumar Yadav, Pradnya Kadam, Unnati Bhalerao, Manisha Tupekar, Dipti Deshpande, Priyanki Shah, LS Shashidhara, Radhika Boargaonkar, Dhawal Patil, Saurabh Kale, Asim Bhalerao, Nidhi Jain, Sanjay Kamble, Syed Dastager, Krishanpal Karmodiya, Mahesh Dharne

## Abstract

The COVID-19 pandemic has emphasized the urgency for rapid public health surveillance methods in early detection and monitoring of the transmission of infectious diseases. The wastewater-based epidemiology (WBE) has emerged as a promising tool to analyze and enumerate the prevalence of infectious pathogens in a population ahead of time. In the present study, real time quantitative polymerase chain reaction (RT-qPCR) and Illumina sequencing was performed to determine the SARS-CoV-2 load trend and dynamics of variants over a longitudinal scale in 442 wastewater (WW) samples collected from 10 sewage treatment plants (STPs) of Pune city, India, during November 2021 to April-2022. In total 426 distinct lineages representing 17 highly transmissible variants of SARS-CoV-2 were identified. The SARS-CoV-2 Omicron variant fragments were detected in WW samples prior to its detection in clinical cases. Moreover, highly contagious sub-lineages of Omicron, such as BA.2.12 (0.8-0.25%), BA.2.38 (0.8-1.0%), BA.2.75 (0.01-0.02%), BA.3 (0.09-6.3%), BA.4 (0.24-0.29%), and XBB (0.01-13.7%) fragments were significantly detected. The longitudinal analysis also suggested the presence of the BA.5 lineage in November 2021, which was not reported in the clinical settings throughout the duration of this study, indicative of silent variant persistence. Overall, the present study demonstrated the practicality of WBE in early detection of SARS CoV-2 variants, which could be useful in tracking future outbreaks of SARS-CoV-2. Such approaches could be implicated in the monitoring of the infectious agents before they appear in clinical cases.

**Highlights:**

□ Omicron fragments were detected in the sewershed samples prior to clinical samples.
□ Omicron sub-lineages BA.2.12, BA.2.38, BA.2.75, BA.3, BA.4, and XBB were prevalent.
□ Lineage composition analysis indicated transition from Delta to Omicron variant indicated cause of third wave in India.
□ Overall, 426 lineages of 17 highly transmissible variants of SARS-CoV-2 were detected in the study.

## 1. Introduction

The SARS-CoV-2 pandemic, initially detected in Wuhan, China in 2019, spreads rapidly all over the world (Global research on coronavirus disease (COVID-19),” W.H.O). As the virus spreads, the emergence of new variants has become a concern for public health officials. The clinical testings have limitations in detecting and tracking the spread of variants (Faria et al., 2021; Li et al., 2020). Also, the SARS-CoV-2 virus epidemic has sparked global efforts to understand its transmission dynamics and the emergence of new variations (Kucirka et al., 2020; Rambaut et al., 2020). To address this challenge, researchers have begun exploring alternative methods for monitoring the spread of SARS-CoV-2, such as wastewater-based epidemiology (WBE) (de Roda Husman et al., 2020; van der Meer et al., 2020).

WBE involves analyzing viral particles present in sewage to infer the spread of infection within a population. WBE has been used to study other viral diseases, such as influenza and hepatitis E (Heijnen and Medema, 2011; Rosa et al., 2014), and has recently been applied to SARS-CoV-2. Studies by Peccia et al., 2020b and Ahmed et al., 2022 have provided essential insights into the feasibility and applicability of WBE for SARS-CoV-2, highlighting its potential as an early detection system for population infection dynamics. Several studies have been conducted globally, with notable contributions from researchers in Germany, Italy, Netherlands, Finland, USA, Japan, France, Canada and India (Maida et al., 2022; Tiwari et al., 2022b; Prado et al., 2021; Stephens et al., 2022; Ho et al., 2022; Amman et al., 2022; Kumar et al., 2020). These studies provides essential information on the detection, quantification and genetic characterization of SARS-CoV-2 in sewage samples (Srivastava et al., 2021; Dharmadhikari et al., 2022; Lamba et al., 2023). The WBE has the potential to detect viral signals earlier than clinical surveillance methods and can offer a less biased assessment of viral heterogeneity and public health (Joshi et al., 2022; Jahn et al., 2022; Morvan et al., 2022). The ongoing COVID-19 outbreak has brought attention to the need for rapid and accurate surveillance of the spread of SARS-CoV-2.

The impact of SARS-CoV-2 has been felt globally since its emergence in 2019 (Chakraborty and Maity, 2020). The virus, which causes COVID-19, has resulted in substantial interruptions to everyday life and economic activities, as well as widespread disease and death. (Shang et al., 2021, Li et al., 2020). One of the critical concerns in the COVID-19 epidemic is the advent of novel SARS-CoV-2 variants, which may be more transmissible and lead to more severe disease (Aleem et al., 2022). Omicron variant is known to be associated with a high number of cases across the globe (Nyberg et al., 2022). The virus has significantly impacted India, with several clinical cases and deaths reported (Ranjan, 2022). Among the Indian cities affected, Pune has recorded one of the highest numbers of SARS-CoV-2 Omicron cases. To comprehend the propagation and evolution of SARS-CoV-2, researchers have used next-generation sequencing (NGS) platforms to analyze genomic sequences of the virus. These studies have shown that different SARS-CoV-2 variants have emerged in vast geographical areas, with varying infectivity and transmission capability levels. In light of this, NGS platforms have proven to be a valuable tool in understanding the evolution of the virus and the dangers it poses to society. Studies by Agrawal et al., 2022 in Germany, Landgraff et al., 2021 in Canada, Wilton, 2021 in London, Crits-Christoph, 2021 in California, and Dharmadhikari et al., 2022 in India have used NGS platforms to identify SARS-CoV-2 mutations in wastewater. These studies have proven that WBE can capture SARS-CoV-2 and its mutants in wastewater, providing insight into the spread of the virus in communities.

The present study is the first in-depth longitudinal study employing wastewater-based SARS-CoV-2 sequencing to delineate the overall dynamics of SARS-CoV-2 lineages in Pune city, India. Here, we utilized WBE to monitor the emergence of SARS-CoV-2 variants, specifically the Omicron lineage in Pune City, analyzing its presence in wastewater samples and assessing its impact on the local population and healthcare system. Understanding the dynamics of the Omicron variant and its sub-lineages will provide insights that can inform public health response and help mitigate the spread of the virus. To achieve our objectives, we systematically analyzed wastewater samples collected from different locations in Pune City, using RT-qPCR and sequencing techniques to detect and quantify the presence of the Omicron variant. This study will provide critical insights into the dynamics of the Omicron variant in Pune City and inform the development of effective public health measures to combat its spread.

## 2. Materials and Methods

### 2.1. Wastewater sample collection and SARS-CoV-2 sequencing

Sewage wastewater samples were taken from the inlets of 10 sewage treatment plants (STPs) in Pune (Fig.1, Table.S1). The sample collection included a grab-sampling method, executed following the CDC, USA guidelines of COVID-19 wastewater sampling, and approved by the Institutional Biosafety Committee (IBSC). Sampling was carried out two times in a week from 10 STPs, in the period of November 2021 - April 2022 in 1L sterile polypropylene bottles and stored at 4°C until processing. The processing was carried out immediately as per CDC, USA guidelines of COVID-19 sewage water sampling. The sampling bottle was wiped with 70% ethanol and subjected to heat inactivation at 60°C. 80 ml of the sample was aliquoted in a 250 ml HDPE bottle (HiMedia, India) and subjected to viral RNA concentration using the polyethylene glycol (PEG) precipitation method. The detailed protocol of sample processing and RT-qPCR is given in supplementary information (Supplementary File.2) Sequencing was carried out at Next Generation Genomics facility at IISER, Pune, India. The library preparation was carried out using Illumina Covid seq RUO kits (Illumina, USA). Sequencing was carried out using Illumina NextSeq 550 sequencer.

**Fig. 1.**
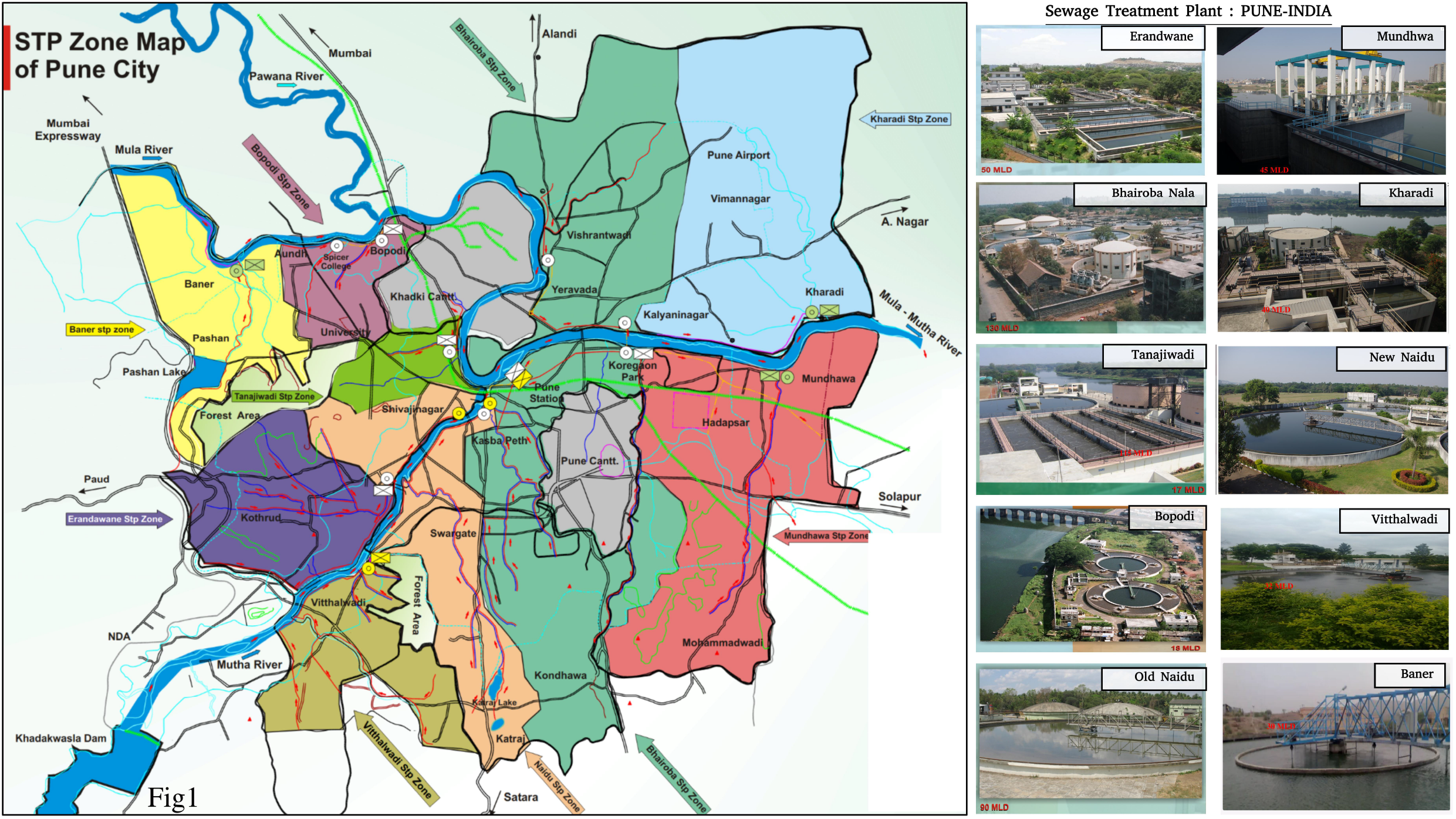
Geographical locations of the sewage treatment facilities sampled in Pune city (India).

### 2.2. Bioinformatics Analysis

In this study, an in-house bioinformatics pipeline was designed to identify the predominant lineages and their signature mutations of SARS-CoV-2 in the sample. The pipeline consisted of several crucial steps to ensure the accuracy of the results. The first step involved quality control and filtering of the raw reads using fastp (Chen et al., 2018), which helps to ensure the quality of the data being analyzed. The filtered reads were then aligned with the reference genome of SARS-CoV-2 (MN908947.3) using BWA-Mem (Li and Durbin, 2010), providing basic alignment and coverage statistics using SAMtools coverage/bedcov (Li et al., 2009). To determine single nucleotide variants (SNVs), iVar (Grubaugh et al., 2019) tool was used with parameters of a minimum base quality score of 20, minimum frequency of 0.03, and minimum read depth of 10. To predict the lineage composition of the SARS-CoV-2 variants in a sample, we utilized LCS (Valieris et al., 2022b), a mixture model designed to determine the variant composition in environmental samples. We used LCS lineage abundance estimates to compute Shannon diversity and Bray-Curtis dissimilarity metrics, as well as an Adonis statistical analysis using R-programming packages such as vegan v2.5-7, ggplot, and phyloseq. Finally, data analysis was performed using scripting in R/Python/Bash to manually check variant-associated mutations and to plot the results.

## 3. Results

### 3.1. Quantification and correlation of SARS-CoV2

We analyzed data collected from 10 sewage treatment plants (STPs) located across the Pune city, covering an estimated 92% of the city’s population between November 2021 and April 2022 (Fig.1, Table.S1). An average of two samples were taken from each location per week to assess the level of community spread. Wastewater samples were processed and analyzed within 24h of sample collection. RT-qPCR was employed to test 442 samples for the presence of SARS-CoV-2 genetic material using three different genetic target regions: N, RdRp, and E gene. A sample was considered positive for SARS-CoV-2 when any two genes showed Ct values lower than 35. In total, 142 out of 442 samples were positive, while the remaining 300 were negative, representing a total positive detection rate of 32.12%. (Table.S2)

To gain insights to the prevalence of SARS-CoV-2 infection in the community, we calculated the amount of virus present in each sample using a viral load calculation tool. The results of this analysis, depicting the levels of SARS-CoV-2 in wastewater and clinically reported cases in Pune (http://cms.unipune.ac.in/~bspujari/Covid19/Pune2/) are illustrated in (Fig.2, Fig.3). A strong correlation for the entire city viral load exhibits a significant association with daily clinically reported COVID-19 positive cases (R^2^=0.81, p<22e-16; Fig.S1.A) and 7-day moving average COVID-19 cases (R^2^=0.83, p<22e-16; Fig.S1.B). The RT-qPCR analysis revealed an increase in viral load during the SARS-CoV-2 surge in January 2022, with the virus detected in the samples throughout the period from November 2021 to April 2022. The viral load was observed lowered in November and December 2021, with an average viral copy of 3.2 gc/ml and 2.2 gc/ml, respectively, while a sharp increase in viral load was noted in January 2023, with an average detection of 83.9 gc/ml. Overall wastewater viral trend forms an early mirror image over clinically reported COVID-19 cases, demonstrating the usefulness of WBE in detecting the early transmission of SARS-CoV-2 infection in the community.

**Fig. 2.**
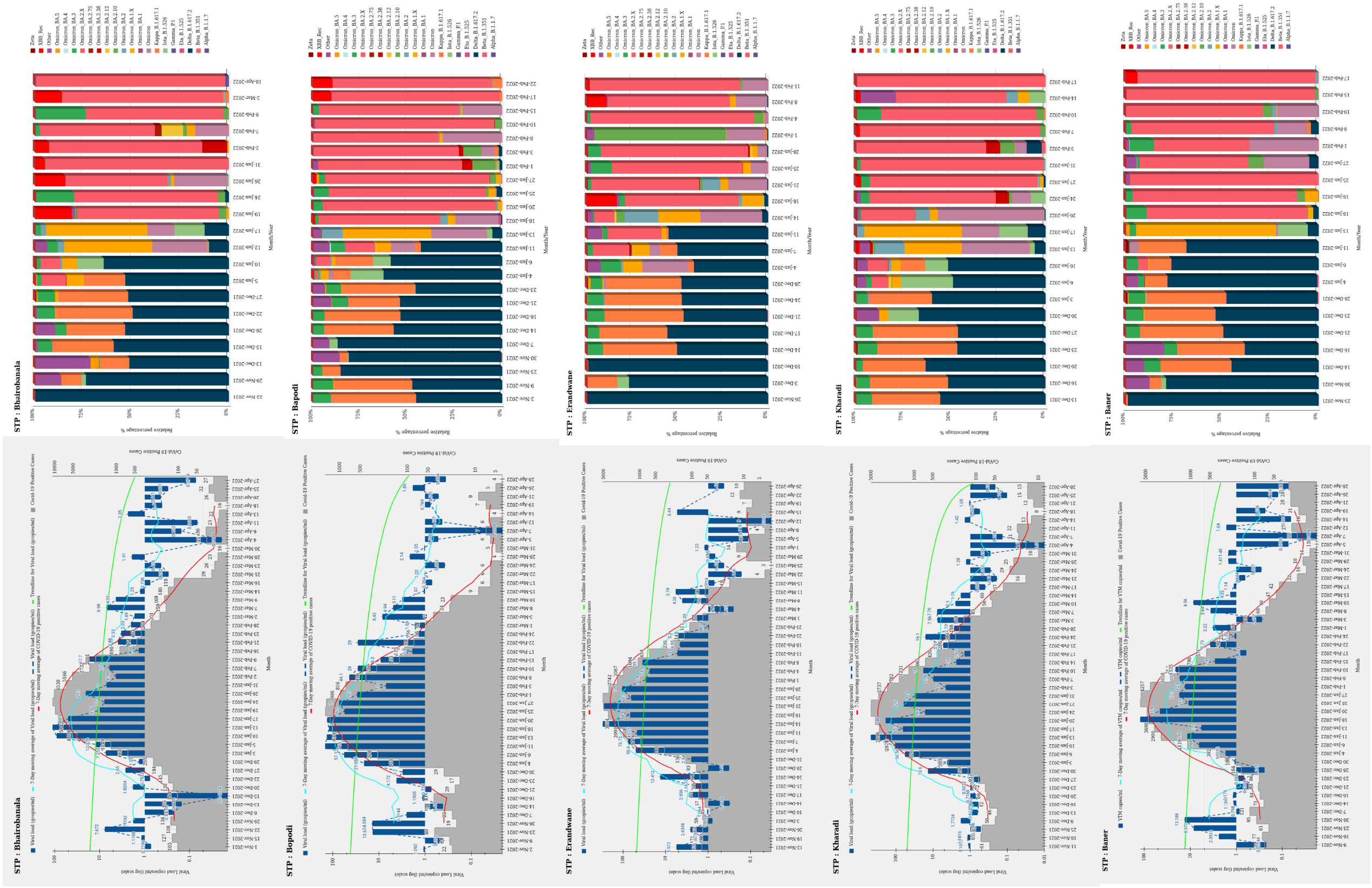
Temporal trends of SARS-CoV-2 viral load in relation to COVID-19 cases and the relative abundance of SARS-CoV-2 variants in sewer shed sites (Bhairobanala, Bopodi, Erandwane, Kharadi, Baner).

**Fig. 3.**
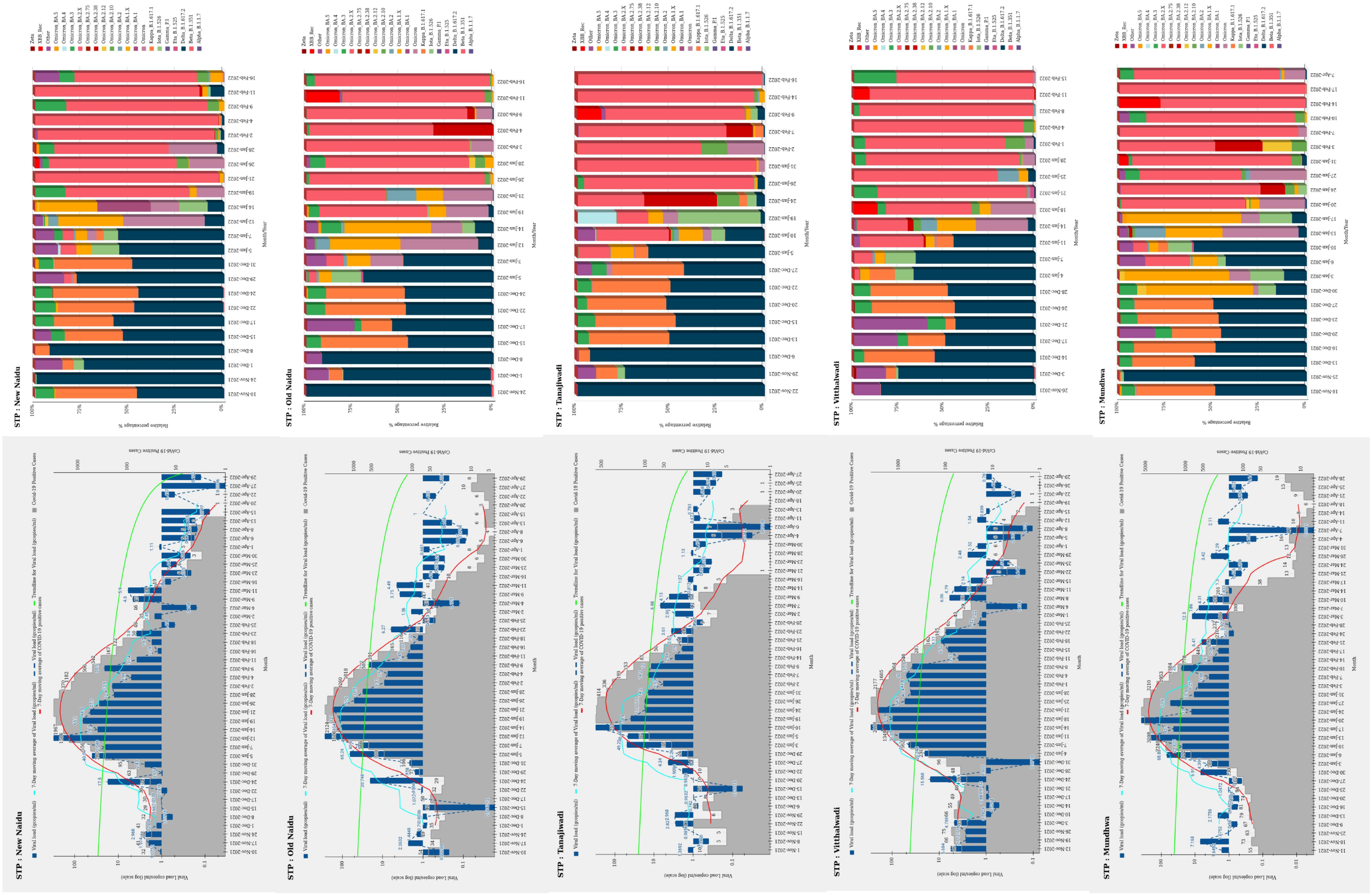
Temporal trends of SARS-CoV-2 viral load in relation to COVID-19 cases and the relative abundance of SARS-CoV-2 variants in sewer shed sites (New Naidu, Old Naidu, Tanajiwadi, Vitthalwadi, Mundhwa).

The SARS-CoV-2 signals were detected across all STPs throughout the study. When COVID-19 entered Pune in its third wave, all STPs began to give early warnings 08 to 17 days before the city’s clinical COVID-19 cases substantially increased. STP sites such as Old Naidu, Mundhwa, and New Naidu and Kharadi gave early warning signals from December 8^th^ to December 16^th^, 2021, sequentially, whereas STP sites like Tanajiwadi, BhairobaNala, Bopodi, and Erandwane began providing early warning signals after December 20^th^, 2021. However, STP sites of Vitthalwadi and Baner gave signals at the same time the clinical cases substantially started increasing (Fig.2, Fig.3), which probably indicates the effective clinical testing in the region. This phenomenon of detecting 08 to 17 days early viral signals from wastewater proves the usefulness of WBE as a system for early detection.

### 3.2. Prediction and diversity of SARS-CoV-2 variants in wastewater

To understand the dynamics of SARS-CoV-2 variants in the city, we conducted a large-scale SARS-CoV-2 sequencing effort on 442 samples collected over six months from November 2021 to April 2022 (Table.S3). A significant proportion of the samples which is 52%, failed to yield sufficient sequencing data due to low viral load as evidenced by high cycle threshold (Ct) values. However, the remaining 48% of samples met our stringent quality criteria, including detection confidence of SARS-CoV-2 and minimum genome coverage of 55%, and were subsequently used for lineage prediction analysis. The study’s findings offer insightful information on the evolution and propagation of SARS-CoV-2 variants within our study population.

The relative frequencies and diversity of SARS-CoV-2 lineages in wastewater were analyzed using the *LCS* tool in a study conducted from November 2021 to April 2022. A total of 426 distinct lineages representing 17 highly transmissible variants of SARS-CoV-2 were identified in WW during this period (Fig.2, Fig.3). The Delta lineage was found to be the most dominant during November and December 2021, with an overall percentage of 80% and 55% respectively (Table.S5), followed by the Kappa lineage, which accounted for 12% and 30% of the total lineages during the same period, while BA.3 and BA.2 lineages accounts for 2.4-6.3% and 0-3% respectively. The BA.1 lineage accounted for only 0.3% and 0.2% in January and February’2022, respectively. The remaining 8% of lineages were classified as “others”. However, small signals of the Omicron lineage started to be detected during November 2021 (2.6%) and December 2021 (7.9%), with a 10-fold increase in January 2022 (70%) to become the most prevalent lineage from January 2022 onwards (Fig.4). Interestingly, the study also detected strong signals of the XBB lineage in March samples accounts for 13.7% to total lineages. This emphasizes the significance of continuing monitoring of SARS-CoV-2 variants in WW to better understand their spread and transmission dynamics. Identifying highly transmissible variants in WW may also affect public health interventions and measures.

**Fig. 4.**
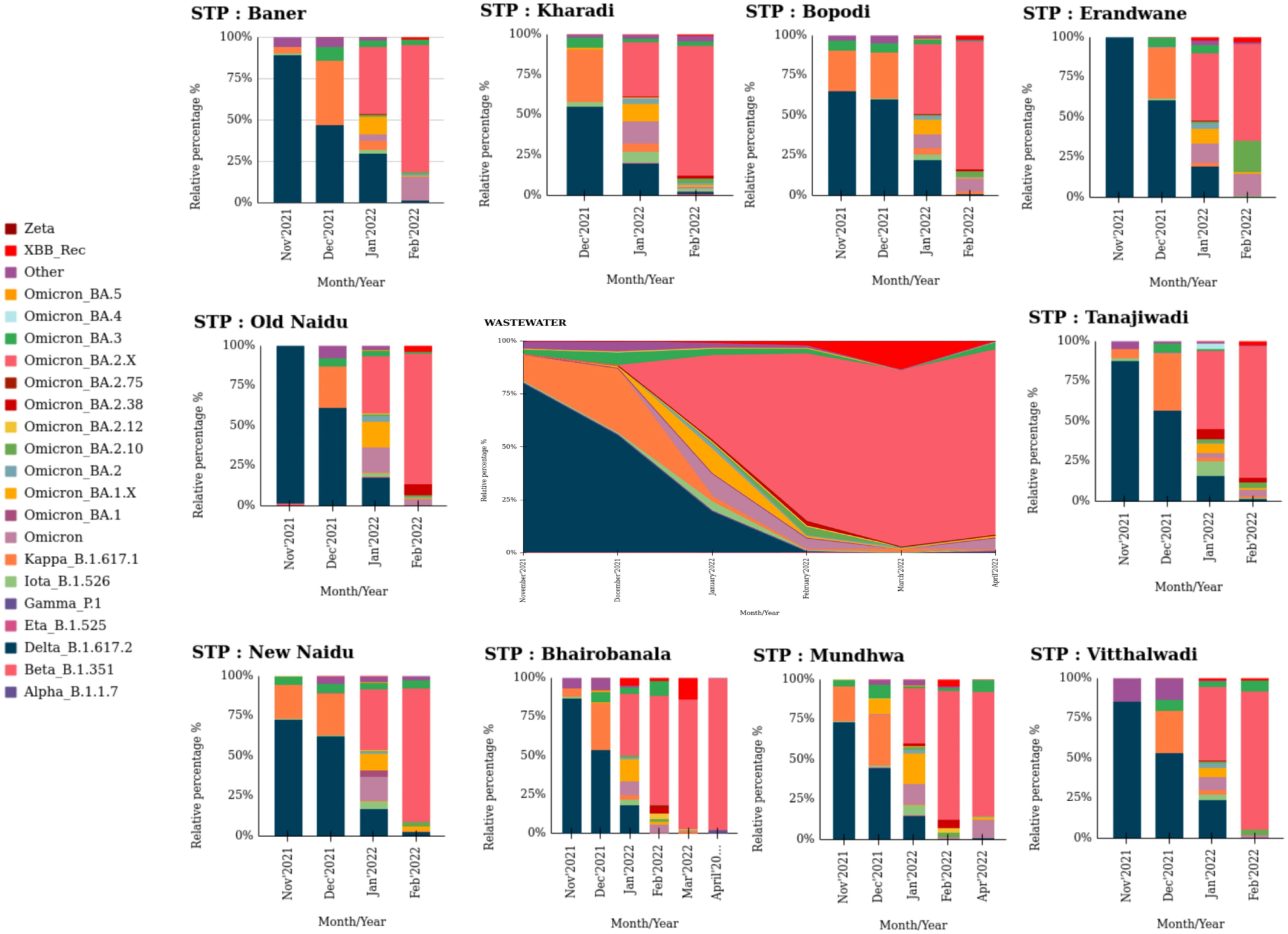
Relative abundance of SARS-CoV-2 lineages in wastewater according to date of collection and sewer shed sites in Pune.

### 3.3. Diversity and correlation between Wastewater and Clinical SARS-CoV-2 data

We aimed to correlate the lineage composition between clinical and wastewater samples of SARS-CoV-2 in Pune city, India. We retrieved 3223 SARS-CoV-2 genomes from GISAID from November 2021 to April 2022, submitted from Pune city (Table.S6). A total of 98 distinct lineages and sub lineages were identified during this period. Based on the predominance of each lineage in India, the lineages were categorized according to the INSACOG categorization for further study (INSACOG Dashboard, Table.S7). The results showed that the overall clinical lineages followed a similar pattern as the wastewater samples. The Omicron lineage and its sub lineages were the dominant lineages from January to April 2022. Omicron BA.2 accounted for 51.6% of all lineages, followed by Omicron BA.2.10 (14.8%), Omicron BA.2.X (13.8%), Omicron BA.1.X (6.3%), Omicron BA.1 (5.6%), and Omicron BA.2.38 (4.9%). (Fig.S3). However, the Delta lineage was the most dominant lineage during November and December 2021. Specifically, the Delta_B.1.617.2 lineage accounted for 88.1% of the total lineages, followed by early signals of Omicron_BA.1.X (5.2%), Omicron_BA.2 (2.8%), and Omicron_BA.1 (2.1%). Similarly, the wastewater samples had dominant lineages of Delta_B.1.617.2 and Kappa during November and December 2021. Later, the Omicron variant takeover the Delta variant and became a dominant variant by January’ 2022 (Fig.4). This transition from Delta to the omicron variant highlights the significance of ongoing monitoring and observation of SARS-CoV-2 lineages in both clinical and wastewater samples to gain a better understanding of their spread and transmission dynamics.

To understand the diversity of SARS-CoV-2 lineages circulating in the community, we calculated Simpson indices for all the samples collected from various sites. These indices measure the total diversity of a community and are commonly used in ecology and epidemiology. The results of our analysis revealed a significant difference in diversity between months (Kruskal-Wallis, p=2.6e-12) but no significant difference by location (Fig.S2.A, Fig.S2.B). To further investigate the variation in diversity across time, we used the Bray-Curtis dissimilarity matrix with the Adonis test to compute beta diversity. This approach allows for comparing community composition between different samples or groups. The analysis found a significant difference in beta diversity across the months (R2 = 0.55, P>0.001) but no significant variation by location (Fig.S2.C, Fig.S2.D). These results suggest that temporal rather than geographic factors primarily influence the diversity of SARS-CoV-2 lineages in Pune. Overall, these findings discern the dynamics of SARS-CoV-2 lineage diversity in the population and highlight the importance of continued monitoring to track emerging variants of concern.

### 3.4. Early detection of Omicron and its sub-lineages

To determine the population’s exposure to SARS-CoV-2 variants, we utilized the *iVar* tool to analyze each sample for co-occurring signature mutations of the Omicron variant (BA.1, BA.2, BA.3, BA.4, BA.5, BA.1.x, BA.2.x, BA.3.x, BA.4.x, BA.5.x). Despite some limitations in the sequencing data due to low RNA concentrations in certain samples, we could still detect characteristic signals of the Omicron variant. Our analysis revealed that wastewater samples taken from the STP at Bhairobanala was the first one to have the Omicron signals : S: D614G, S: H655Y, S: N679K, S: P681H, N: P13L) and Tanajiwadi (S: N440K, S: S477N, S: T478K, S: E484A, S: Q493R, S: Q498R, S: N501Y, S: Y505H, S: T547K, S: D614G, E: T9I, on November 1st, 2021, before it was classified as a VOC by the World Health Organization (“Tracking SARS-CoV-2 variants,” WHO) on November 26th, 2021 (Table.S8). The Omicron was also detected in samples collected from the STP at Bopodi ((S: L452R, S: T478K, E: T9I) on November 2nd, 2021, before it was confirmed in Pune’s first clinical case on December 5th, 2021. Furthermore, our analysis revealed that Omicron and its sub lineages were detected in wastewater samples collected from nine out of ten STPs in Pune, indicating widespread community exposure to this variant before its detection in clinical cases (Table.S9). These findings emphasize the use of wastewater-based surveillance in tracking the emergence and spread of SARS-CoV-2 variants in the community. It is important to note that our investigation was carried out in India, and the Omicron variant was discovered in Botswana on November 2011. As such, our data suggest that Omicron may have been circulating in wastewater prior to its detection in clinical cases. The findings of this investigation offer crucial insights into the early detection and spread of SARS-CoV-2 in the community and support the implementation of WBE as a complementary approach to traditional clinical surveillance.

Moreover, to evaluate the rise of variation in the lineage of SARS-CoV-2 in the population, we employed a linear mixed model analysis to examine the correlation between the initial detection of variant of concern in wastewater and the date when clinical samples were collected from Pune city and submitted to GISAID. Our examination revealed a trend of increasing presence of the Omicron variant (B.1.1.529) and its sub lineages (BA.1, BA.2, BA.2.12, BA.2.X, BA.2.38, BA.3, BA.4, BA.5) starting from January 2022. As time progressed, however, we noted a reduction in the frequency of the Kappa (B.1.617.1) and Delta (B.1.617.2) lineages, with the Omicron variant completely dominating by February 2022 (Fig.5). Additionally, we observed an upward trend in the prevalence of the XBB lineage starting from January 2022. These findings emphasize the crucial importance of ongoing monitoring of the variations in SARS-CoV-2 in order to inform public health strategies.

**Fig. 5.**
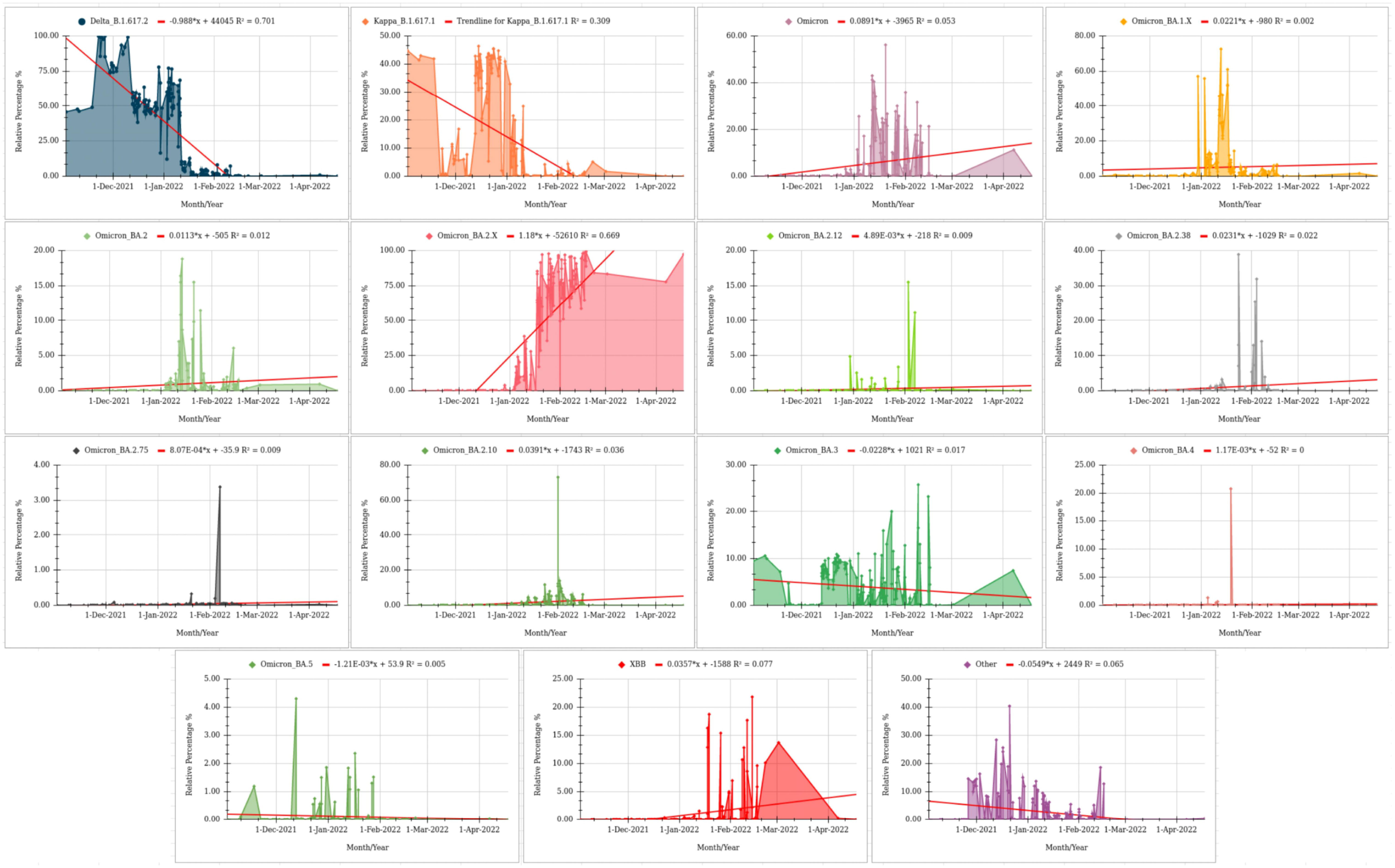
Temporal trends of dominant variants of SARS-CoV-2 in wastewater samples between November 2021 and April 2022.

## 4. Discussion

The emergence of new SARS-CoV-2 variants has been a key cause of concern in the ongoing COVID-19 epidemic. One such variant, Omicron, has been identified in India (Gao et al., 2021). This variant has raised concerns due to its high transmissibility and potential to evade immunity from previous infections or vaccines (Shrestha et al., 2021; Zhang, 2022; Yadav and Kumar, 2022). Our study utilizes WBE to identify, track, and assess the SARS-CoV-2 genetic variants in Pune city, India. We collected 442 wastewater samples from 10 locations, representing an estimated 7.4 million individuals. WBE has been used in several studies globally to identify and quantify the SARS-CoV-2 fragments in wastewater (Srivastava et al., 2021; Dharmadhikari et al., 2022; Joshi et al., 2022; Jahn et al., 2022; Morvan et al., 2022). By applying this model at the city level, we aim to prevent the spread of COVID-19 and better understand the dynamics of SARS-CoV-2 transmission in Pune city.

In this study, we used RT-qPCR and Illumina sequencing to quantify and monitor the genetic characteristics of SARS-CoV-2 variants. Our findings show a strong correlation between the viral load and clinical cases of COVID-19. Additionally, our data suggest that a significant amount of viral shedding likely occurs early in infection, particularly from asymptomatic individuals, before they seek medical attention and are tested for Covid-19 (Lamba et al., 2023). The SARS-CoV-2 persists in faeces samples longer than in respiratory and serum samples (Prakash et al., 2021). In the present study, the SARS-CoV-2 was detected before an increase in clinical cases highlighting the potential utility of WBE as a tool for early detection and prevention of SARS-CoV-2 transmission. By monitoring STPs, public health officials can identify outbreaks and implement strict management strategies at the earliest stages of severe disease.

The SARS-CoV-2 genetic material was quantified using RT-qPCR, followed by Illumina sequencing to determine the trends of SARS-CoV-2 lineages disseminated in the community. Detecting SARS-CoV-2 titers in wastewater is challenging amid the fragmentation and PCR inhibitors, making it difficult to quantify and sequence for estimating lineage/variant composition (Baaijens et al., 2021). However, in this study, LCS (Lineage decomposition for SARS-CoV-2), a model for predicting the lineage composition in a sample, was utilized, which captures and classifies variant-specific mutations (Valieris et al., 2022). Our results showed that relative to the clinical testing, SARS-CoV-2 RNA content in wastewater provides a more accurate indicator for local infection dynamics (Karthikeyan, 2021); Randazzo et al., 2020; Polo et al., 2020). We found a wide range of SARS-CoV-2 lineages in wastewater, with some lineages that infect a substantial proportion of people exhibiting similar patterns. Additionally, we discovered low prevalent mixed lineages in wastewater that were undetectable from clinical data, suggesting that such lineages infected only a small number of individuals.

Furthermore, weak signals from novel variants imply the arrival of a new lineage into the population that could not be emphasized in clinical data-indicating the limited testing or testing for symptomatic individuals exclusively. We discovered early XBB signals and other Omicron sub-lineages that were not previously reported in clinical data. This discrepancy highlights the lack of required sequencing, i.e. at least 0.5% of the positive cases (Grimaldi et al., 2022), which impacts the ability to detect SARS-CoV-2 variants quickly and undermines the global pandemic preparedness. Our research emphasizes the effects of socioeconomic disparities, population density, and wastewater treatment as powerful strategies to stop the spread of SARS-CoV-2 (Sharifi and Khavarian-Garmsir, 2020). It is imperative to continue monitoring and testing wastewater for SARS-CoV-2 to prevent the spread of new variants and to ensure global pandemic preparedness. We aimed to quantify the SARS-CoV-2 genetic material in wastewater samples and determine the genomic trends of SARS-CoV-2 lineages circulating in the community. Our results showed that wastewater samples exhibit greater alpha diversity than clinical samples. Additionally, the viral composition of the city did not show regional variability, as the beta diversity of SARS-CoV-2 did not differ by STP locations. However, there was a significant variation (35%) across clinical and wastewater samples, and distinct and common clusters of SARS-CoV-2 lineages were formed over time, suggesting introduction and elimination of various lineages in the community over time (Karthikeyan et al., 2022; Gu et al., 2022). These findings emphasize the significance of wastewater surveillance as a complementary tool to clinical testing in understanding the dynamics of SARS-CoV-2 transmission in the community.

We investigated the presence of different lineages and sub-lineages of SARS-CoV-2 in wastewater samples collected from Pune, India. Our findings showed the presence of several variants of the Omicron lineage, which have been linked to increased transmissibility, adverse health impacts, and hospitalization (Khandia et al., 2022; Araf et al., 2022.). Importantly, our results indicated that three out of the ten catchment areas had detectable levels of Omicron in the wastewater samples before the first clinical case of Omicron-positive in Botswana on November 11th, 2021: 10 days earlier in Bhairobanala, 9 days earlier in Bopodi, and 10 days earlier in Tanajiwadi. Additionally, nine out of the ten catchment areas had Omicron signals in the wastewater samples before the first reported clinical case of Omicron in Pune city on December 5th, 2021: 19 days earlier in Baner, 34 days earlier in Bhairobanala, 33 days earlier in Bopodi, 23 days earlier in Erandwane, 24 days earlier in Kharadi, 18 days earlier in New Naidu, 25 days earlier in old Naidu, 34 days earlier in Tanajiwadi, and 24 days earlier in Mundhwa. This implies that a substantial portion of the population was infected and shedding the virus before being diagnosed through clinical testing (He et al., 2020).

Furthermore, we discovered six highly transmissible lineages of Omicron in the wastewater samples before their detection in clinical samples from the city: BA.2.12, BA.2.38, BA.2.75, BA.3, BA.4, and XBB detected 20, 8, 153, 242, 127, and 279 days earlier, respectively. Additionally, we found weak indications of lineage BA.5 in November 2022, which has yet to be reported in any clinical cases from Pune city. Our findings demonstrate the utility of WBE for detecting and monitoring the presence of SARS-CoV-2 variants in the community. Testing of wastewater captures the combined contributions of every individual in a catchment region, providing a less biased representation of viral diversity and public health (Karthikeyan et al., 2022). In contrast, delays in detection from clinical samples may be due to limited testing and sequencing or asymptomatic or home-testing cases (Mercer and Salit, 2021). Additionally, our study showed that sequencing data from wastewater samples could be used to monitor the prevalence of variants and estimate their growth rate, using significantly fewer samples than required for clinical samples. Overall, these results substantiates the importance of WBE as a complementary approach to traditional clinical surveillance for detecting and monitoring the spread of SARS-CoV-2 variants. Furthermore, it also highlights the potential of WBE to detect the emergence of new variants before they are identified in clinical samples.

## 5. Conclusion

The present study highlighted the efficacy of wastewater-based epidemiology (WBE) as a formidable tool in the early warning, detection, and monitoring of SARS-CoV-2 variants. By conducting a comprehensive analysis of wastewater samples collected between November 2021 and April 2022, we could detect the Omicron variant fragments before clinical cases, providing valuable insights for public health officials to implement preventative measures. Our findings highlight the potential of WBE as a practical approach for real-time surveillance of infectious outbreaks, not only for SARS-CoV-2 but also for other pathogens. Integrating WBE into existing public health surveillance methods can significantly enhance our ability to understand the transmission dynamics and evolution of infectious diseases and, ultimately, help in preventing future outbreaks.

## Data Availability

All data produced in the present study are available upon reasonable request to the authors

## Abbreviations

WW: Wastewater
COVID-19: coronavirus disease 19
SARS-CoV-2: severe acute respiratory syndrome coronavirus 2
WHO: The World Health Organization
VOI: variants of interest
VOC: variants of concern
WBE: wastewater-based epidemiology
E: envelope E
N: nucleocapsid
WWTP: wastewater treatment plants
PEG-8000: polyethylene glycol-8000
IISER: Indian Institute of Science Education and Research
NCL: National Chemical Laboratory

## Data availability

All data produced in the present study are available upon reasonable request to the authors. Clinical COVID-19 data was used from CMS dashboard (http://cms.unipune.ac.in/~bspujari/Covid19/Pune2/)

## Acknowledgments

Authors wish to thank Directors of CSIR-NCL, IISER-Pune, and PKC for encouragement and support. We gratefully acknowledge Pune Municipal Corporation (PMC) for permission to carry out the sampling and use clinical COVID-19 data. Authors are indebted to Prof. Aurnab Ghose, IISER-Pune (for grant administration support and discussions), Dr. Dayanand Panse-ESF (for support and pre-preparations in sampling logistics) and Ms. Rachel Samson (for processing and RNA extractions of the subset samples). Authors also acknowledge https://biorender.com/ for creation of images. The manuscript has been thoroughly checked for plagiarism content using iThenticate software.

