## Supplementary material for "Genomic surveillance reveals early detection and transition of Delta to Omicron Lineages of SARS-CoV-2 Variants in wastewater treatment plants of Pune, India": Supplemantry_File_2

**Supplementary File.1.**

**Materials and Methods**

**Wastewater sample collection and processing**

Briefly, the samples were centrifuged at 5000xg (5910 R, Eppendorf, Germany) for 10 minutes at 4°C followed by successive filtration of supernatant twice with Whatman qualitative filter paper grade 1 (Merck, USA), followed by filtration with 0.22 µm PES filter membrane (diameter 47mm) using vacuum filtration assembly (Tarsons, India). The resulting filtrate was added to a 250ml HDPE bottle (HiMedia, India), containing 8% (w/v) PEG 8000 (HiMedia, India) and 1.7% of molecular biology grade NaCl (w/v) (HiMedia, India) and was incubated for 2 h at 4°C in a rotating shaker water bath (Equitron medica, India) at 175 rpm. The mixture was centrifuged at 10000xg for 1h at 4°C to collect the viral pellet. The pellet was resuspended in 280 μL TE buffer (pH 7.0). Extraction of Viral RNA was carried out using QIAmp Viral RNA Minikit (Qiagen, Germany) according to the manufacturer’s instructions. The final elution of RNA was done in 80 µL of elution buffer and stored at -80°C for future use.

**Viral Reverse Transcriptase-quantitative PCR assay (RT-qPCR)**

The SARS-CoV-2 was detected using RY-qPCR using Genepath Dx kit (ICMR approved) as per the manufacturer’s instructions on 7500 Fast Real-Time PCR system (Applied Biosystems, USA). RNA-dependent polymerase (RdRP) gene, nucleocapsid (N) genes, and Envelop (E) gene were used for quantitative detection of SARS-CoV-2 RNA in the samples. At least two out of three genes need to be present in the detectable range (Ct<35) then the sample will be considered positive. The Covid-19 Viral Load Calculation Tool was used to quantify the SARS-CoV-2 viral load in the samples (RUO).

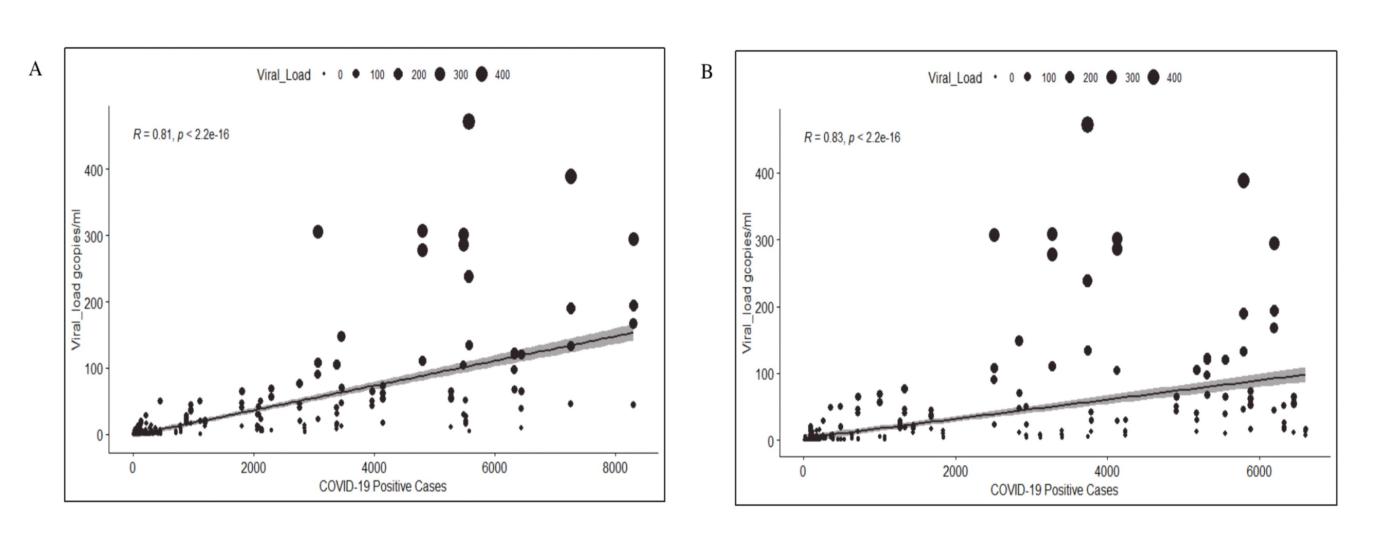

**Fig.S1** Linear Regression between Viral RNA concentration and Clinical Cases. **A.)** Correlation between Viral RNA concentration and per day COVID-19 cases. **B.)** Correlation between Viral RNA concentration and 7-day moving average clinical cases. The linear regression fitting is shown by the solid black line. 95% confidence interval from the fitting standard error is indicated by the light gray region.

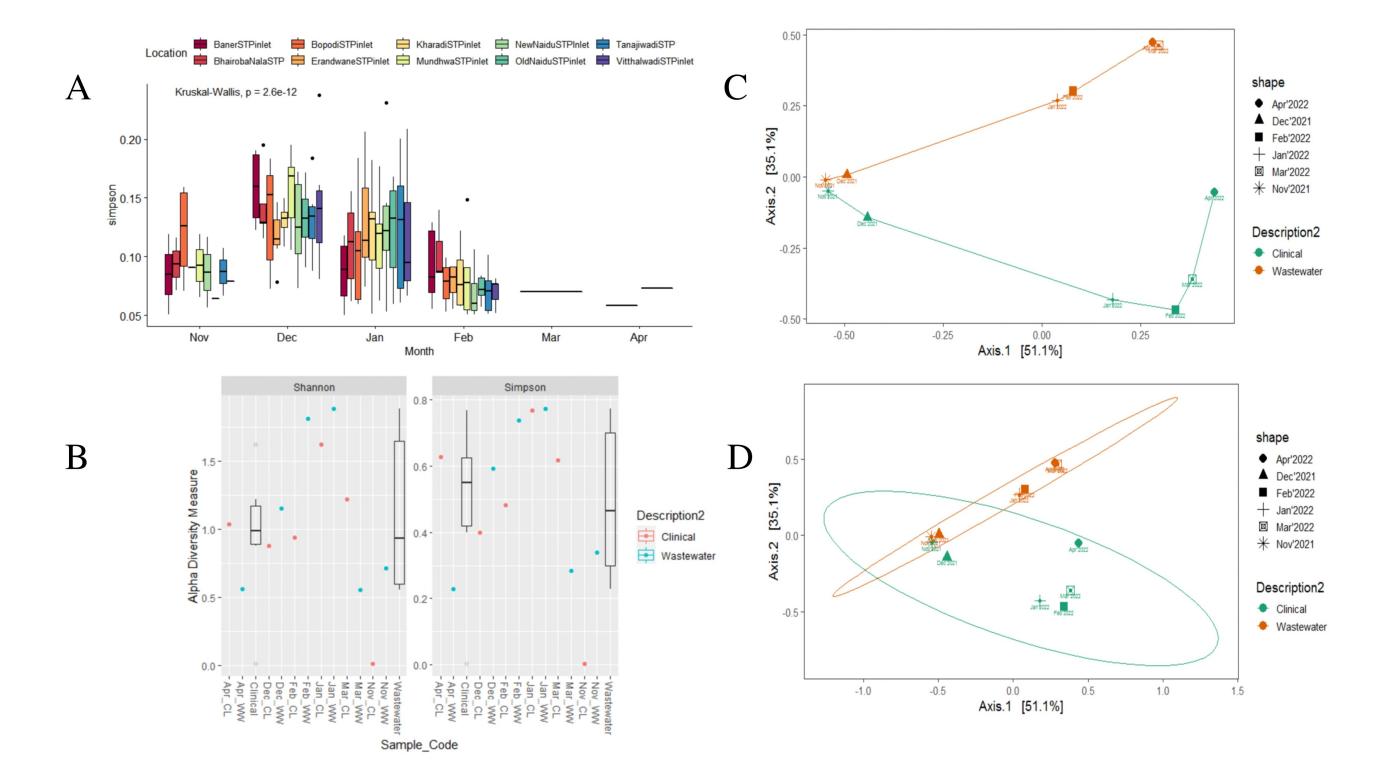

**Fig.S2** SARS-CoV-2 lineage diversity. **A.**) Simpson diversity analysis (alpha diversity) of SARS-CoV-2 lineages across months. **B.)** Simpson and Shannon diversity analysis among lineages wastewater and clinical. **CandD.)** Beta diversity analysis (Bray–Curtis dissimilarities) among clinical and wastewater lineages by month.

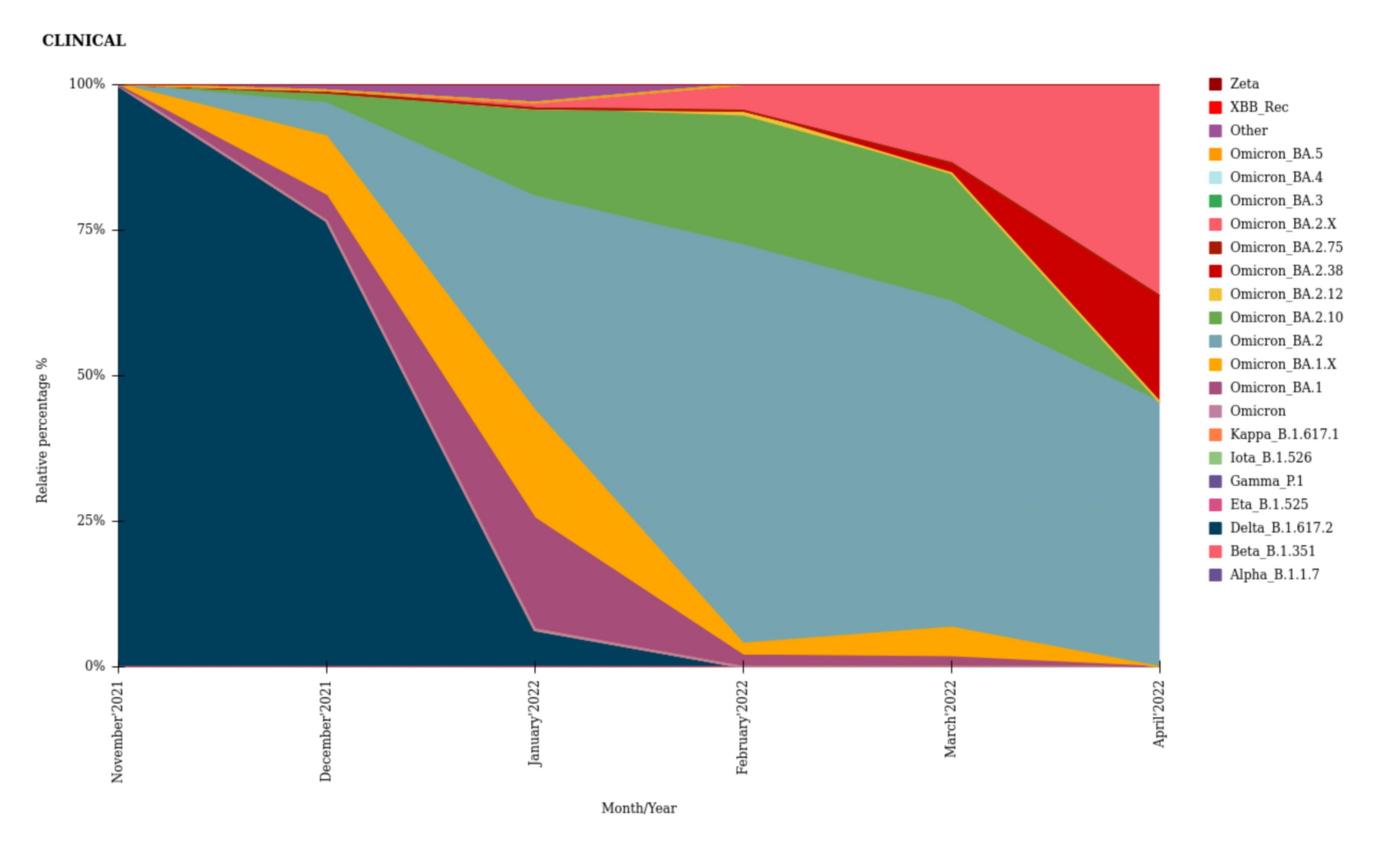

**Fig.S2** Relative abundance of SARS-CoV-2 lineages in clinical by date of collection in Pune.
